# Similar viral loads in Omicron infections regardless of vaccination status

**DOI:** 10.1101/2022.04.19.22274005

**Authors:** Yosuke Hirotsu, Makoto Maejima, Masahiro Shibusawa, Yume Natori, Yuki Nagakubo, Kazuhiro Hosaka, Hitomi Sueki, Hitoshi Mochizuki, Toshiharu Tsutsui, Yumiko Kakizaki, Yoshihiro Miyashita, Masao Omata

## Abstract

**Background:** Although SARS-CoV-2 booster vaccinations are underway, breakthrough infections with Omicron variants are occurring. This study analyzed associations between Omicron sublineage (BA.1.1 and BA.2) viral load and vaccination history.

**Methods:** Viral loads in nasopharyngeal swabs were evaluated by quantitative real-time PCR, and the virus strain was evaluated by whole-genome analysis or TaqMan assay.

**Results:** A total of 611 patients positive for an Omicron SARS-CoV-2 variant were included; 199 were unvaccinated, 370 had received two vaccine doses, and 42 had received three doses. Similar viral loads and Ct values of BA.1.1 and BA.2 were detected regardless of vaccination history. No correlations between age and BA.1.1 and BA.2 viral load were observed.

**Conclusion:** Omicron-infected patients who had received a third vaccine dose had viral loads similar to patients with two doses or who were unvaccinated.

## Introduction

The outbreak of severe acute respiratory syndrome coronavirus 2 (SARS-CoV-2) has caused an estimated 497 million cases of coronavirus disease 2019 (COVID-19) and 6.18 million deaths worldwide. SARS-CoV-2 has acquired mutations throughout its evolution, driving the emergence of new viral strains. To date, five variants of concern have been designated by the World Health Organization: Alpha, Beta, Gamma, Delta, and Omicron. Omicron emerged in South Africa in 2021 and has spread worldwide. Unlike other variants of concern, the Omicron variant has approximately 30 mutations in its spike protein. Some of its mutations (e.g., 69-70del, T95I, G142D/143-145del, K417N, T478K, N501Y, N655Y, N679K, and P681H) are also present in the Alpha, Beta, Gamma, and/or Delta variants. These mutations lead to increased transmissibility, increased viral binding affinity, and increased immune escape [1].

Several Omicron sublineages have been reported; BA.1 and BA.1.1 were initially most prevalent, but BA.2 is now predominating. Antibody activity against BA.1/BA.1.1 and BA.2 differs because the viruses have different mutations in the spike protein [2, 3]. A booster dose restores effectiveness in preventing infection and reduces disease severity [4-6]. Nevertheless, breakthrough infections with Omicron variants have been reported after a third dose [7, 8]. Thus, it is important to understand the virus levels that are typically present after breakthrough infection to better evaluate infection control, quarantine, and public health measures. However, the viral loads in cases of Omicron breakthrough infection after booster vaccination are not fully clear.

Patients infected with Omicron BA.1.1 or BA.2 were included in this study regardless of vaccination history, and nasopharyngeal swabs were used to measure viral loads.

## Methods

### SARS-CoV-2 diagnostic testing

We performed SARS-CoV-2 diagnostic testing on samples collected from January 10, 2022 to April 7, 2022. The following diagnostic testing platforms were used in this study: COVID-19 reverse transcription-PCR performed in accordance with the protocol developed by the National Institute of Infectious Diseases in Japan [9], the FilmArray Respiratory Panel 2.1 test performed with the FilmArray Torch system (bioMérieux, Marcy-l’Etoile, France) [10], the Xpert Xpress SARS-CoV-2 test performed on a Cepheid GeneXpert system (Cepheid, Sunnyvale, CA, USA) [11], and the Lumipulse antigen test performed on a LUMIPULSE G600II system (Fujirebio, Inc., Tokyo, Japan) [12, 13]. All tests were conducted on material obtained from nasopharyngeal swabs immersed in viral transport media (Copan, Murrieta, CA, USA).

### Quantitative reverse transcription-PCR (RT-qPCR)

To detect SARS-CoV-2, we performed one-step RT-qPCR to amplify the nucleocapsid (N) gene of SARS-CoV-2 as previously described [14]. The human ribonuclease P protein subunit p30 (*RPP30*) gene was used as the internal positive control (Integrated DNA Technologies, Coralville, IA, USA) [14]. The RT-qPCR assays were performed on a StepOnePlus Real-Time PCR system (Thermo Fisher Scientific, Waltham, MA, USA) with the following cycling conditions: reverse transcription at 50 °C for 5 min, inactivation of reverse transcription at 95 °C for 20 s, followed by 45 cycles of denaturation at 95 °C for 3 s and annealing/extension at 60 °C for 30 s. The threshold was set at 0.2. In accordance with the national protocol (v. 2.9.1) [9], samples were determined to be positive if a visible amplification plot was observed and negative if no amplification was observed.

### SARS-CoV-2 genome analysis

Whole-genome sequencing was conducted in accordance with a previously described method using the nasopharyngeal swab samples collected from SARS-CoV-2-positive patients. In brief, SARS-CoV-2 genomic RNA was reverse-transcribed into cDNA and amplified using the Ion AmpliSeq SARS-CoV-2 Research Panel or the Ion AmpliSeq SARS-CoV-2 Insight Research Assay (Thermo Fisher Scientific) on the Ion Torrent Genexus system in accordance with the manufacturer’s instructions [15-17]. The sequencing reads were processed and their quality assessed using Genexus software with SARS-CoV-2 plugins. The sequencing reads were then mapped and aligned using the torrent mapping alignment program. After initial mapping, a variant call was performed using the Torrent Variant Caller. The COVID19AnnotateSnpEff plugin was used to annotate the variants. Assembly was performed using Iterative Refinement Meta-Assembler [18].

The viral clade and lineage classifications were conducted using Nextstrain [19] and Phylogenetic Assignment of Named Global Outbreak Lineages (PANGOLIN) [20]. The sequence data were deposited in the Global Initiative on Sharing Avian Influenza Data (GISAID) EpiCoV database [21].

### TaqMan assay

We used the pre-designed TaqMan SARS-CoV-2 Mutation Panel to detect SARS-CoV-2 spike Δ69–70, G339D, L452R, and Q493R (Thermo Fisher Scientific) in SARS-CoV-2-positive samples [22]. The TaqMan MGB probe for the wild-type allele was labeled with VIC dye, and the probe for the variant allele was labeled with FAM dye. This TaqMan probe system can detect both wild-type and variant sequences of SARS-CoV-2. TaqPath 1-Step RT-qPCR Master Mix CG was used as master mix. Real-time PCR was conducted on a Step-One Plus Real-Time PCR system (Thermo Fisher Scientific).

### Statistical analysis

All statistical tests and visualizations were performed with R (v4.1.1) or RStudio (http://www.r-project.org/). Plotting in R also made use of the ggplot2 (v3.3.5), dplyr (v1.0.7), tidyr (v1.1.3), patchwork (v1.1.1), gtsummary (v1.5.2), and flextable (v.0.7.0) packages. Bartlett’s test was used to test for equality of variances. Statistical analyses (Kruskal–Wallis rank–sum test; Pearson’s chi-square test; Fisher’s exact test, pairwise t-test with Holm correction, Pearson’s product–moment correlation coefficient) were conducted. P-values < 0.05 were considered statistically significant.

## Results

### Vaccination history and Omicron infection

This study included 611 patients who were diagnosed as SARS-CoV-2-positive between January 10, 2022 and April 7, 2022. The vaccination history of the patients was as follows: 199 were unvaccinated, 370 had received two doses of vaccine, and 42 had received three doses of vaccine. The median ages were 14 years (IQR: 19, 60) for the unvaccinated group, 44 years (IQR: 28, 68) for the two-dose group, and 48 years (IQR: 34, 70) for the three-dose group (Table 1, p < 0.001). The vaccination status by age was consistent with the priority given to older adults to receive a third dose. No significant differences were found between vaccination history and sex (p = 0.7) or disease severity (p = 0.2) (Table 1).

**Table 1.** Patient characteristics.

| Characteristic | Overall,<br>n = 611 | Unvaccinated,<br>n = 199 | 2nd vaccination,<br>n = 370 | 3rd vaccination,<br>n = 42 | p-value<sup>1</sup> |
| --- | --- | --- | --- | --- | --- |
| Age, median (IQR) | 37 (19, 60) | 14 (5, 40) | 44 (28, 68) | 48 (34, 70) | **<0.001** |
| Sex, n (%) |  |  |  |  | 0.7 |
| Female | 290 (47%) | 96 (48%) | 172 (46%) | 22 (52%) |  |
| Male | 321 (53%) | 103 (52%) | 198 (54%) | 20 (48%) |  |
| Symptom, n (%) |  |  |  |  | 0.2 |
| Mild | 155 (76%) | 27 (68%) | 120 (80%) | 8 (57%) |  |
| Moderate I | 25 (12%) | 7 (18%) | 15 (10%) | 3 (21%) |  |
| Moderate II | 22 (11%) | 5 (12%) | 14 (9.3%) | 3 (21%) |  |
| Severe | 2 (1.0%) | 1 (2.5%) | 1 (0.7%) | 0 (0%) |  |
| Unknown<sup>2</sup> | 407 | 159 | 220 | 28 |  |
| Lineage, n (%) |  |  |  |  | **<0.001** |
| BA.1.1 | 453 (74%) | 136 (68%) | 299 (81%) | 18 (43%) |  |
| BA.2 | 158 (26%) | 63 (32%) | 71 (19%) | 24 (57%) |  |

<sup>1</sup> Kruskal–Wallis rank–sum test; Pearson's chi-square test; Fisher's exact test
<sup>2</sup>Symptoms were not assessed because the individual was an outpatient.

Whole-genome sequencing or TaqMan analysis was performed to identify Omicron sublineages. Of the 611 patients, 453 had BA.1.1 and 158 had BA.2. In those who had received a third dose, BA.2 (15.2%, 24/158) was more often detected than BA.1.1 (4.0%, 18/453). These results indicate that BA.2 is better able than BA.1.1 to cause breakthrough infection in those who have received a booster dose.

### Vaccination history and viral load in patients infected with Omicron

To analyze whether vaccination history altered the viral load after infection with the Omicron sublineages, we quantitatively assessed viral load in nasopharyngeal swabs by RT-qPCR (Figure 1). In BA.1.1-infected individuals, the mean viral loads ± SD were 5.3 ± 1.4 (range: −0.9–7.8) in the unvaccinated group, 5.4 ± 1.5 (−0.3–7.7) in the two-dose group, and 4.6 ± 1.7 (2.2–6.8) in the three-dose group, while they were 5.4 ± 1.6 (range: 1.0–7.7), 5.8 ± 1.5 (−0.4–8.2), and 5.5 ± 1.7 (0.5–7.9), respectively, in the BA.2-infected individuals (Figure 1A). In BA.1.1-infected individuals, the mean cycle thresholds (Ct) ± SD were 20.5 ± 5.1 (range: 12–44) in the unvaccinated group, 20.2 ± 5.1 (12–38) in the two-dose group, and 22.5 ± 5.7 (15–31) in the three-dose group, while they were 19.9 ± 5.6 (range: 12–35), 18.4 ± 5.0 (11–37), and 19.9 ± 6.0 (12–38), respectively, in the BA.2-infected individuals (Figure 1B). No significant differences in viral load or Ct values were found among the groups with different vaccination histories (p > 0.05, pairwise t-test with Holm correction).

**Figure 1.**
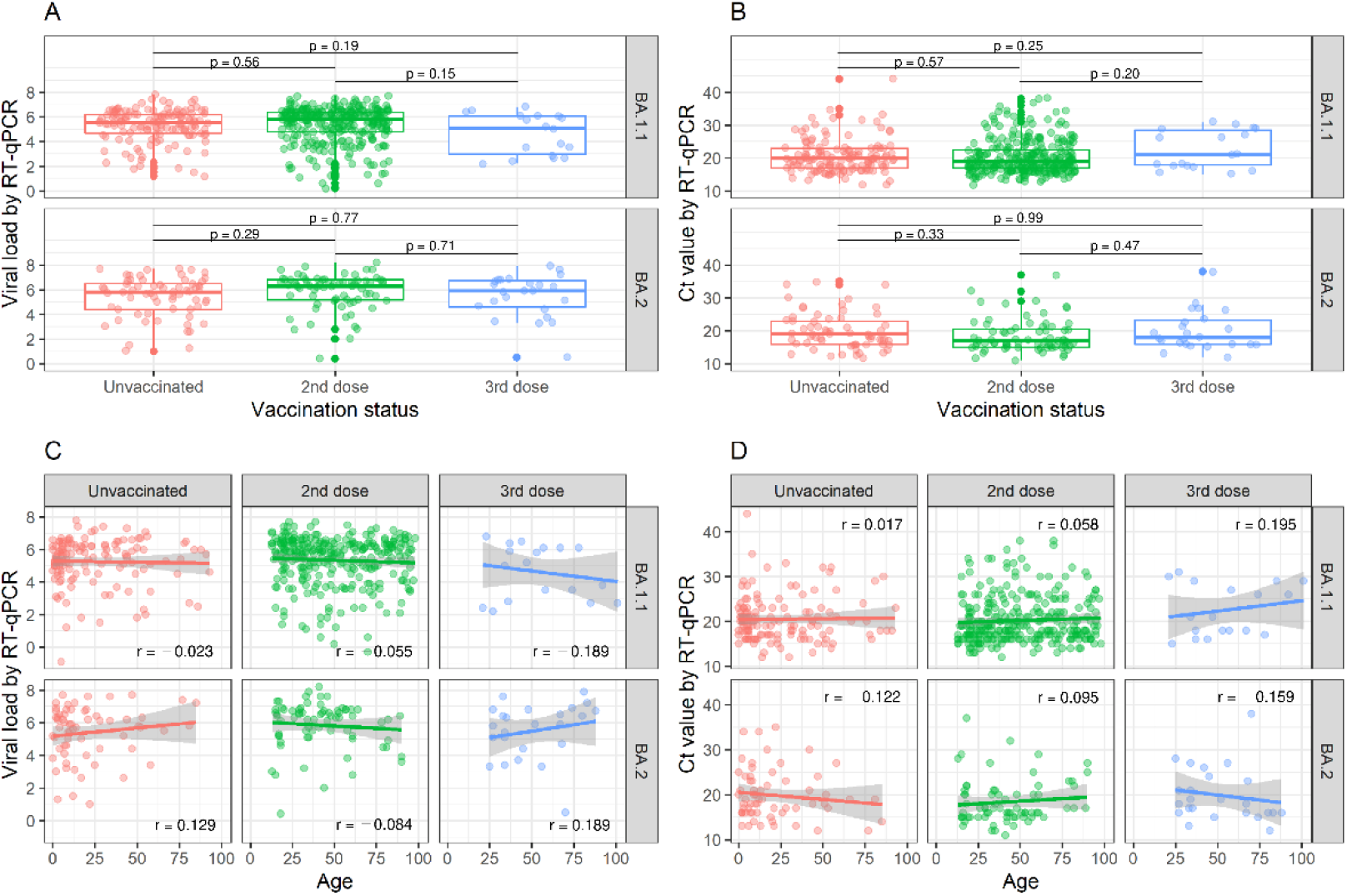
Omicron viral load according to vaccination status. **(A, B)** Box plots show viral load (A) or Ct value (B) in BA.1.11- and BA.2-infected individuals according to vaccination status: unvaccinated, 2^nd^ dose, or 3^rd^ dose. The viral loads and Ct values were determined by RT-qPCR. Groups were compared by pairwise t-tests. All p-values were >0.05. **(C, D)** Correlation plots show the association between age and viral load (C) or Ct value (D) in individuals infected with BA.1.11 or BA.2. Pearson’s correlation coefficients (r) were all less than 0.2, indicating no correlation. The gray regression line background indicates the 95% confidence interval.

We next analyzed whether the viral load after vaccination was correlated with age (Figures 1C and 1D). For those with BA.1.1 infection, the correlation coefficients between age and viral load were r = −0.023 (p = 0.80) for the unvaccinated, r = −0.055 (p = 0.35) for the two-dose group, and r = −0.189 (p = 0.45) for the three-dose group, while for those with BA.2 infection they were r = 0.129 (p = 0.31), r = −0.084 (p = 0.49), and r = 0.189 (p = 0.38), respectively (Figure 1C). For those with BA.1.1 infection, the correlation coefficients between age and Ct value were r = 0.017 (p = 0.85) for the unvaccinated, r = 0.058 (p = 0.32) for the two-dose group, and r = 0.195 (p = 0.44) for the three-dose group, while for those with BA.2 infection they were r = −0.122 (p = 0.34), r = 0.095 (p = 0.43), and r = −0.159 (p = 0.46), respectively (Figures 1D). There was no correlation between age and the viral load, regardless of vaccination history (p > 0.05, Pearson’s correlation coefficient), indicating that when breakthrough infection occurs, the virus can achieve a high viral load regardless of vaccination history and age.

## Discussion

We analyzed the amount of virus present in nasopharyngeal swab samples from patients who became infected even after receiving a third vaccine dose. These patients showed similar amounts of virus as unvaccinated patients. This was consistent with our finding that patients produced a similar amount of virus regardless of age. To our knowledge, this is the first study of viral loads in patients infected with the Omicron BA.1.1 and BA.2 sublineages post vaccination. The findings indicate the potential for secondary transmission from infected individuals even after they have received a booster vaccination. Therefore, non-pharmaceutical interventions, such as mask-wearing and physical distancing, are necessary to prevent the transmission of Omicron variants from both vaccinated and unvaccinated individuals.

The rapid spread of the Omicron variant worldwide has led to its replacement of the Delta variant [21]. Omicron has multiple mutations in the spike protein, raising concerns that antibodies may be less effective than they were against the ancestral strain and other variants of concern [23-25]. Booster vaccination can lower the risk of infection with Omicron, but the protection weakens over time [4, 6]. Infections have been reported despite high levels of anti-spike antibodies after the third vaccination [7]. Our data indicate the need to maintain non-pharmaceutical measures (e.g., mask use, ventilation, physical distancing) to suppress the spread of infection because high viral loads were found even after individuals had received a booster vaccination.

This study had limitations. First, it did not assess the amount of culturable, viable virus, so it is unknown whether the detections represented infectious virus. Second, it did not take into account the time since vaccination, and there may have been variation in antibody titers. Third, there may be bias because the number of vaccine doses differed with age. Therefore, further analysis with a larger sample size is needed.

In summary, individuals infected with the Omicron variant might still be able to produce infectious virus even if they have received a booster vaccination. Therefore, isolating patients with a breakthrough infection could help mitigate the spread of SARS-CoV-2.

## Data Availability

All data produced in the present study are available upon reasonable request to the authors

## Acknowledgments

We thank all medical and ancillary hospital staff for their support. We also thank Katherine Thieltges from Edanz (https://jp.edanz.com/ac) for editing a draft of this manuscript.

